# Several countries in one: a mathematical modeling analysis for COVID-19 in inner Brazil

**DOI:** 10.1101/2020.04.23.20077438

**Authors:** G.B. de Almeida, T.N. Vilches, C.P. Ferreira, C.M.C.B. Fortaleza

## Abstract

Early 2020 and the world experiences its very first pandemic of globalized era. A novel coronavirus, SARS-Cov-2, is the causative agent of severe pneumonia and rapidly spread through many nations, crashing health systems. In Brazil, the emergence of local epidemics in major metropolitan areas is a concern. In a huge and heterogeneous country, with regional disparities and climate diversity, several factors can modulate the dynamics of COVID-19. What should be the scenario for an inner Brazil and what can we do to control infection transmission in each one of these locations? In this paper, a mathematical model was developed to simulate disease transmission among individuals in several scenarios, differing by the intensity and type of control measures. Mitigation strategies rely on social distancing of all individuals, and detection and isolation of infected ones. The model shows that control effort varies among cities. The social distancing is the most efficient method to control disease transmission but improving detection and isolation of infected individuals can help loosening this mitigation strategy.

## Introduction

It has been few months since the first confirmed case of a novel coronavirus pneumonia in Wuhan, China, and now the world experiences its very first pandemic on the globalized era. SARS-Cov-2 rapidly spread through the current connected continents, and the World Health Organization has declared a health emergency on international concern, which made many countries taking serious mitigation and suppression strategies^1^.

These strategies take importance when we look at the epidemics dynamics. The first studies estimated that the basic reproductive number of COVID-19 was 2.68 (95% CrI 2,47-2,86)^2^,3, which means one infected person can spread the virus to almost three people in a totally susceptible community. As there is no treatment or vaccines available, the best way to control the virus was diminishing the social contact. China has shown the world that when people stay at home, the virus circulation is controlled and we have more time for preparing the health systems, producing individual protection equipment, developing research and minimizing the consequences^4^.

In Brazil, the introduction of COVID-19 happened later than in many other locations and that gave us time to analyse all the new scientific evidences and the control measures taken overseas^5^. However, a country with continental dimensions cannot work with a single plan response. São Paulo is the biggest metropolitan area of South America where the epidemic has grown in an important way the last few weeks^6^. However, what is going to be the scenario for an inner Brazil and what can we do on infection control in each one of these locations?

Mathematical modeling has taken great importance when applied into epidemics^7^,8. Since the earliest population studies on plague or measles, the methods were refined. Today, with the parameters well established and the most sensitive parameterizations, such as contact patterns matrices, we can estimate with a good chance of accuracy how an epidemic will behave in a specific population and what should be our immediate response to the problem^9^.

Mathematical models may allow us to draw best and worst scenarios for a COVID-19 epidemic situation in small and medium cities in inner Brazil^10^. Our main objective is to study how the disease might behave in specific cities of the country and see what happens when we combine strategies: diminishing social contact plus testing and isolating positive cases. The consequences of relaxing restrictions is a theme of debate now, and in Brazil it is happening before the peak of epidemic has occurred, while the number of cases still growing over the country. Mask wearing, mass testing, early detection of imported cases and monitoring of effective reproduction number are strategies that have been discussed and adopted around the world^11^.

## Results

For all the cities chosen, the mathematical model was run. We assume that temperature and humidity may influence the virus spread^12^. The effect of temperature (*T*) and humidity (*H*) on *R*_0_ was considered using the expression given by^13^ *R*_0_ = 3.314 *−* 0.0225*T −*0.0158*H*. Data from temperature and humidity for each city is showed in Table 1 together with the calculated *R*_0_ value. We obtained 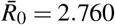 with minimum and maximum values as 2.678 and 2.928 for Mossoró-RN and Lages-SC. Mossoró is situated on Northern Brazil, where the climate is humid and hot. Lages, however, belongs to the brazilian south region, where, in April, the predominance is cold and dry days. Several other abiotic factors can modulate disease transmission but the influence of it on *R*_0_ is not established yet^14^. The range of *R*_0_ is in agreement of what has been observed in Brazil and others countries for COVID-19. In particular, until now, the number of confirmed cases in Santa Catarina and Rio Grande do Norte, respectively where Mossoró and Lages are located, are 596 and 1025.

**Table 1.**
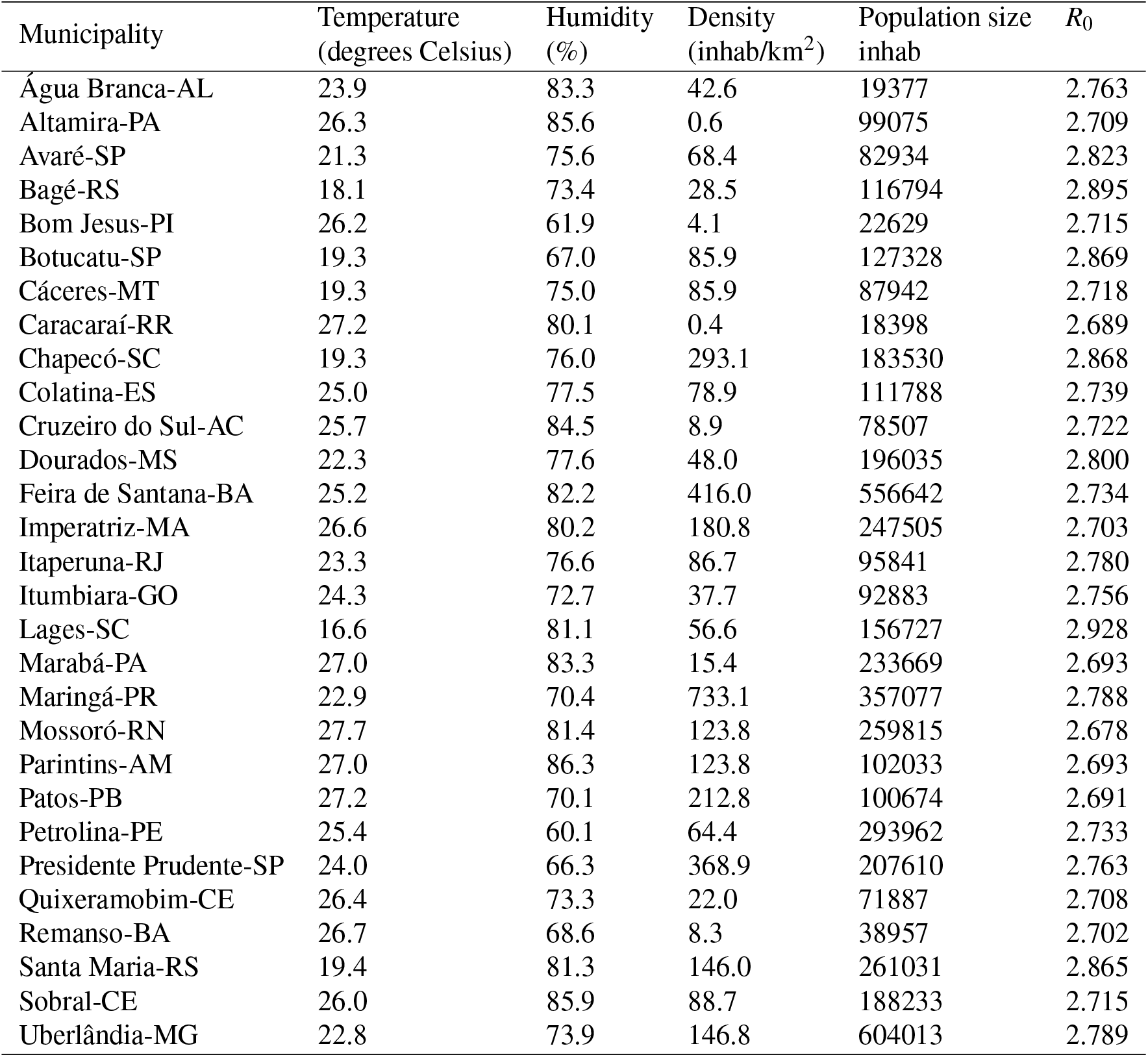
Municipalities and their characteristic abiotic and biotic factors that may modulate Covid-19 transmission.

**Table 2.** Parameters of the model and their values.

| Parameter | Description | Value |
| --- | --- | --- |
| $\mu$ | mortality rate | $1/75 \text{ years}^{-1}$ |
| $\sigma$ | additional mortality rate | $[0, 0.02]$ |
| $\alpha$ | transition rate among age classes | $1/5 \text{ years}^{-1}$ |
| $\eta^{-1}$ | latent period | 3 days |
| $\gamma^{-1}$ | infectious period | 6.4 days |
| $\tau^{-1}$ | isolation period | $\{1, 2, 5, 6\} \text{ days}$ |
| $\varepsilon$ | detection and isolation rate | $1/3 \text{ days}^{-1}$ |
| $\psi$ | fraction of infected that are detected | $[0, 1]$ |
| $\xi, \nu$ | reduction on the infection transmission | $[0, 1]$ |

We calculated the number of cumulative cases per 10000 inhabitants for each one of the municipalities and ranked them, from the least to the most infected. For this analysis, no control was considered. In Figure 3a, we have all infected individuals and only infected individuals older than 50 years. In Figure 3b, we also ranked the proportion of lethal cases per 10000 inhabitants, i.e., the number of deaths divided by the number of cases. In all cases, we considered late detection of infected individuals.

**Figure 1.**
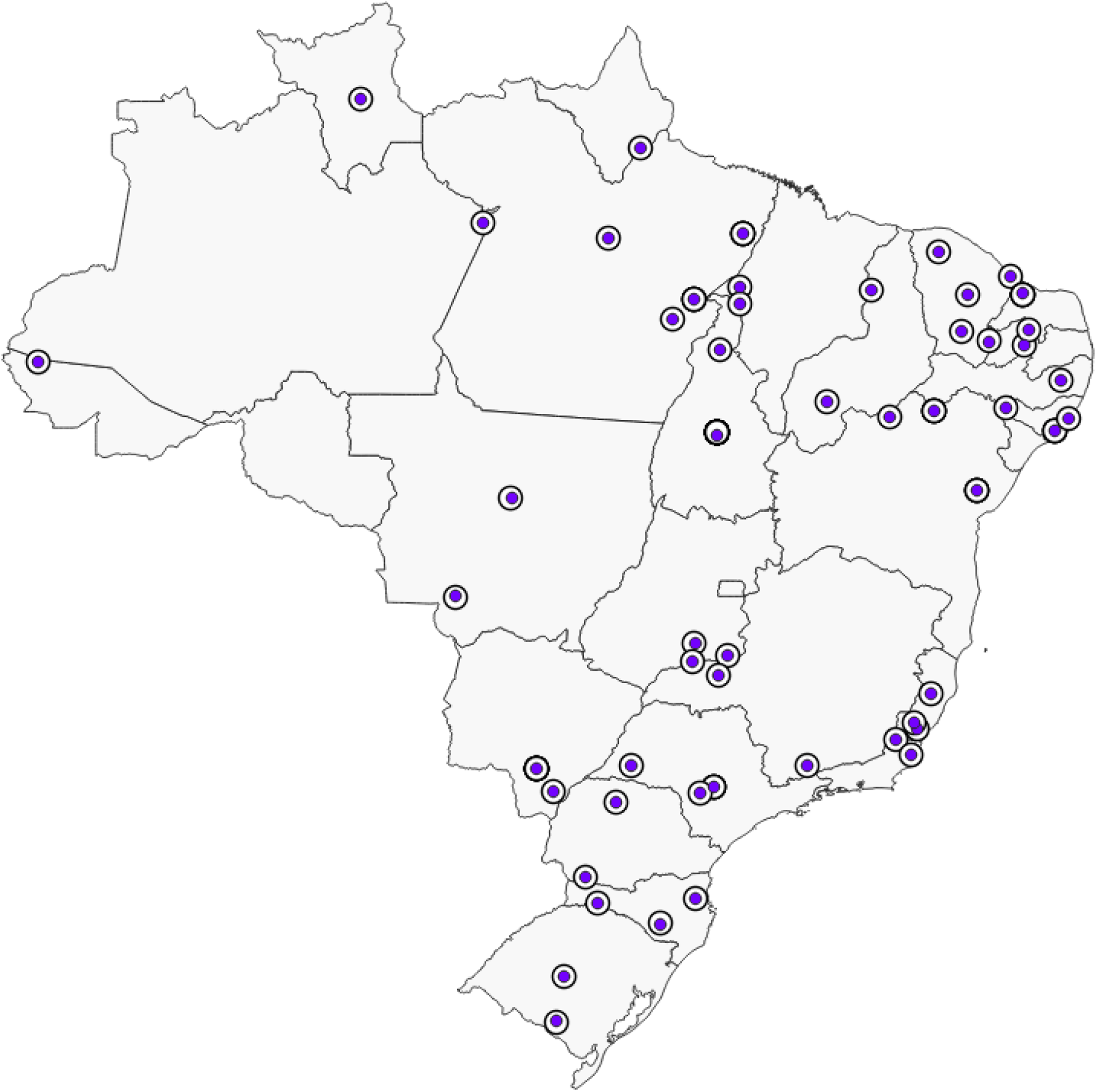
Geographic localization of each chosen municipalities to be evaluated in the present study.

**Figure 2.**
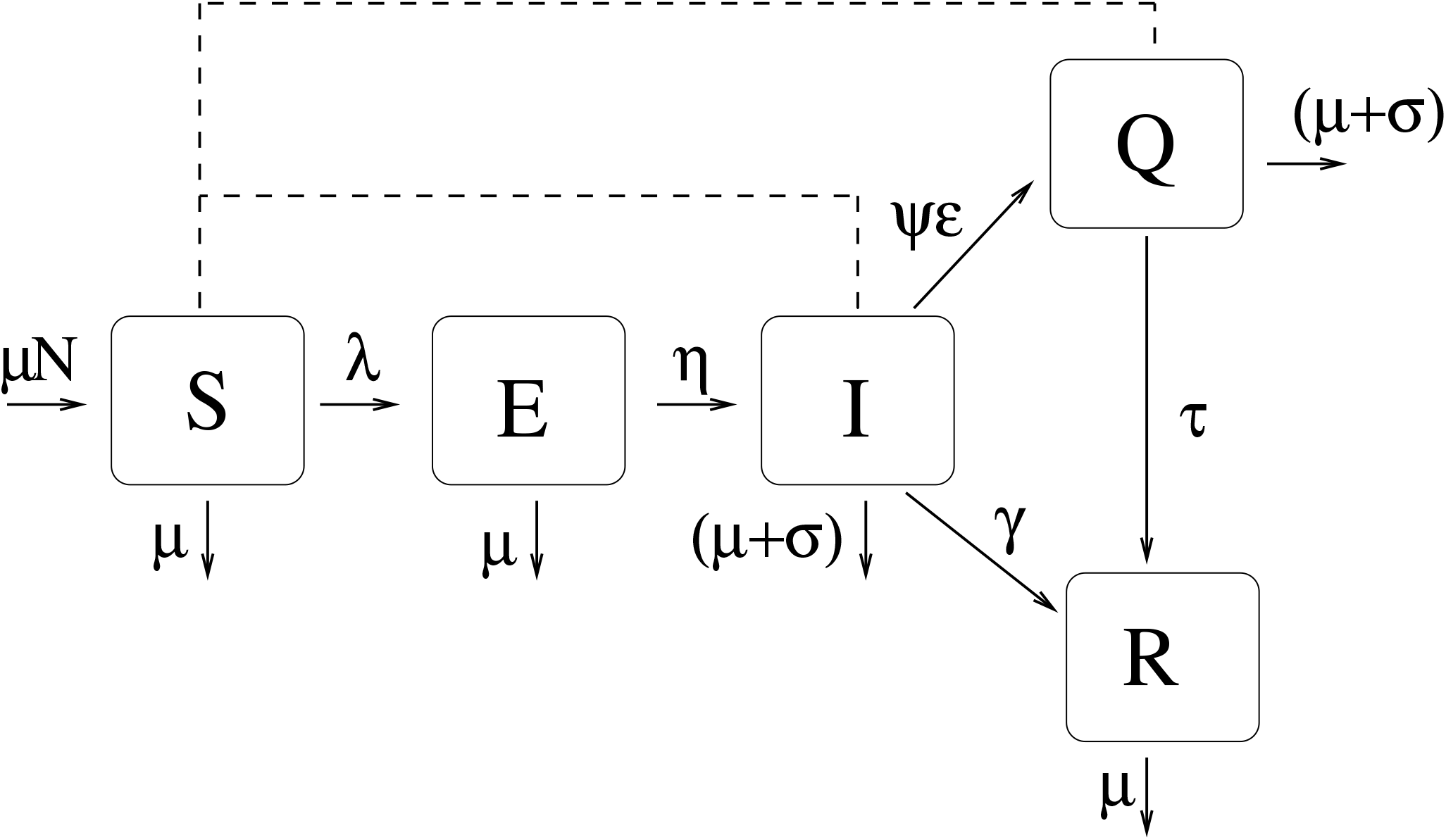
The variables of the model are susceptible (*S*), exposed (*E*), Infected(*I*), Isolated (*Q*) and Recovered individuals (*R*). The continuous line indicates transitions between compartiments and the dashed line indicates interactions between compartiments that contributes to the infection force, *λ*. The parameters are *µ, σ, η*^*−*1^, *γ*^*−*1^, *τ*^*−*1^, *ε*, and *ψ*, respectively, mortality/natality rates, additional mortality rate, latent period, infectious period, isolation period (while infectious), rate at which individuals are detected and isolated, and the fraction of infected individual that are detected.

**Figure 3.**
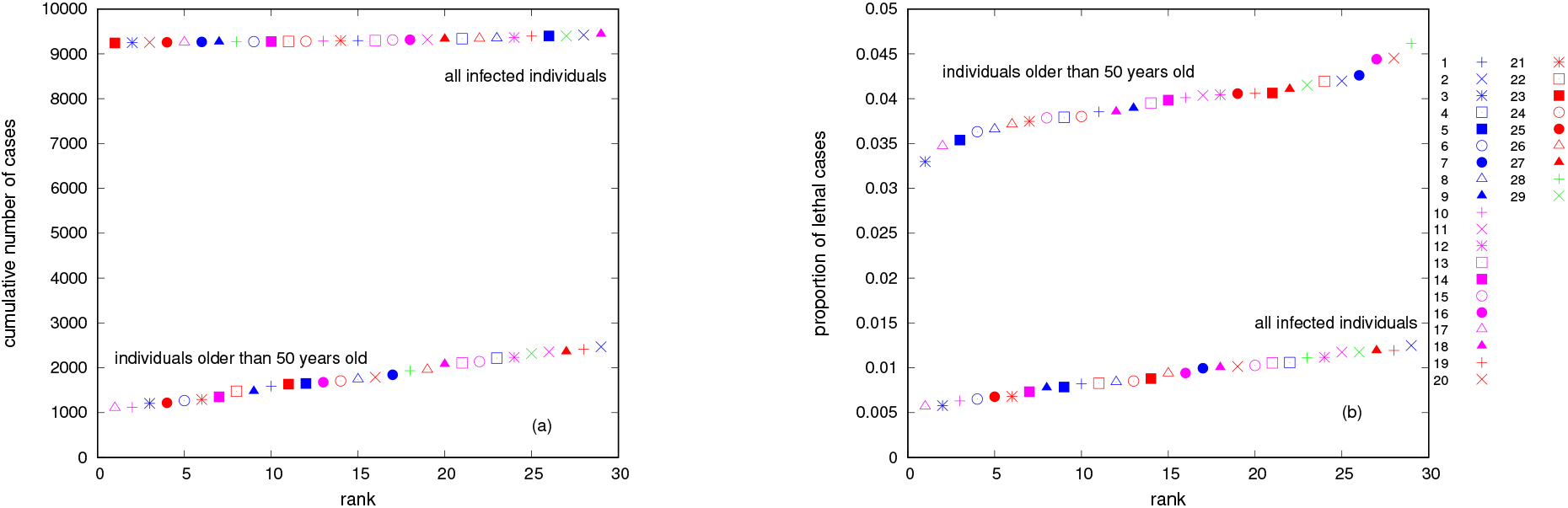
In (a) we have the cumulative number of cases per 10000 inhab versus city rank from the least infected to the most infected one, and in (b) the proportion of lethal cases versus city rank. In both panel we present the sum over all infected individuals and over individuals older than 50 years old. The cities are identified by (1) Feira de Santana, (2) Bagé, (3) Caracaraí, (4) Maringá, (5) Chapecó, (6) Altamira, (7) Remanso, (8) Dourados, (9) Imperatriz, (10) Cruzeiro do Sul, (11) Presidente Prudente, (12) Avaré, (13) Colatina, (14) Bom Jesus, (15) Itumbiara, (16) Agua Branca, (17) Marabá, (18) Lages (19) Santa Maria, (20) Patos, (21) Petrolina, (22) Sobral, (23) Mossoró, (24) Cáceres, (25) Parintins, (26) Uberlândia, (27) Itaperuna, (28) Quixeramobim, (29) Botucatu.

Figure 4 shows the percentage of reduction on the number of cases versus the reduction on contact rate, (1 *− ξ*). In green, blue, red, and orange we have, respectively, Maringá-PR, Lages-SC, Caracaraí-RR, and Patos-PB. The two panels were done for different values of *ψ*, where *ψ* is the fraction of population tested, in (a) we have *ψ* = 0.1 and in (b) *ψ* = 0.3. The vertical lines highlight the efficacy of disease control when the reduction on contact rate is set up to 50%, 60% and 70%.

**Figure 4.**
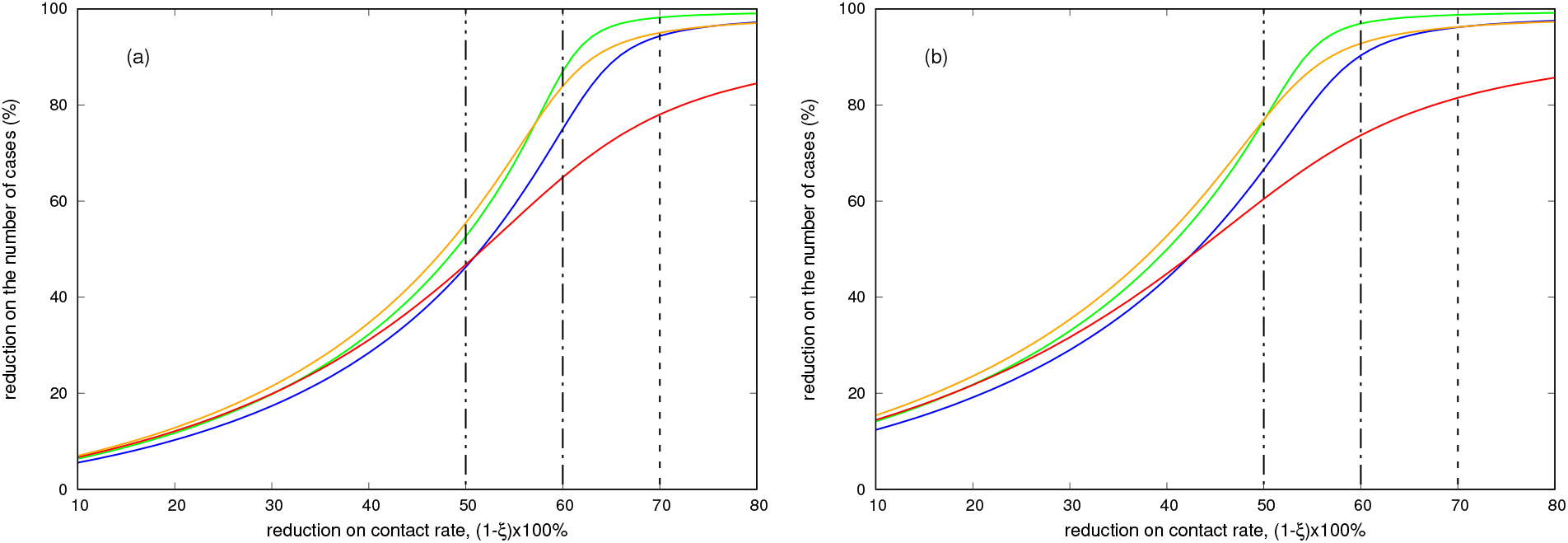
Reduction on the number of cases versus reduction on the contact rate, 1 *− ξ*. In green we have Maringá (*R*_0_ = 2.787), in blue Lages (*R*_0_ = 2.927), in red Caracaraí (*R*_0_ = 2.689), and in orange Patos (*R*_0_ = 2.691). In (a) *ψ* = 0.1 and in (b) *ψ* = 0.3, where *ψ* is the fraction of population tested.

Figure 5 shows the reduction on the number of cases versus the time of starting control, *t*_*s*_. The parameters are the same as in Figure 4. To achieve at least 70% of reduction on the number of cases control has to start before thirty days after the introduction of the first infected individuals.

**Figure 5.**
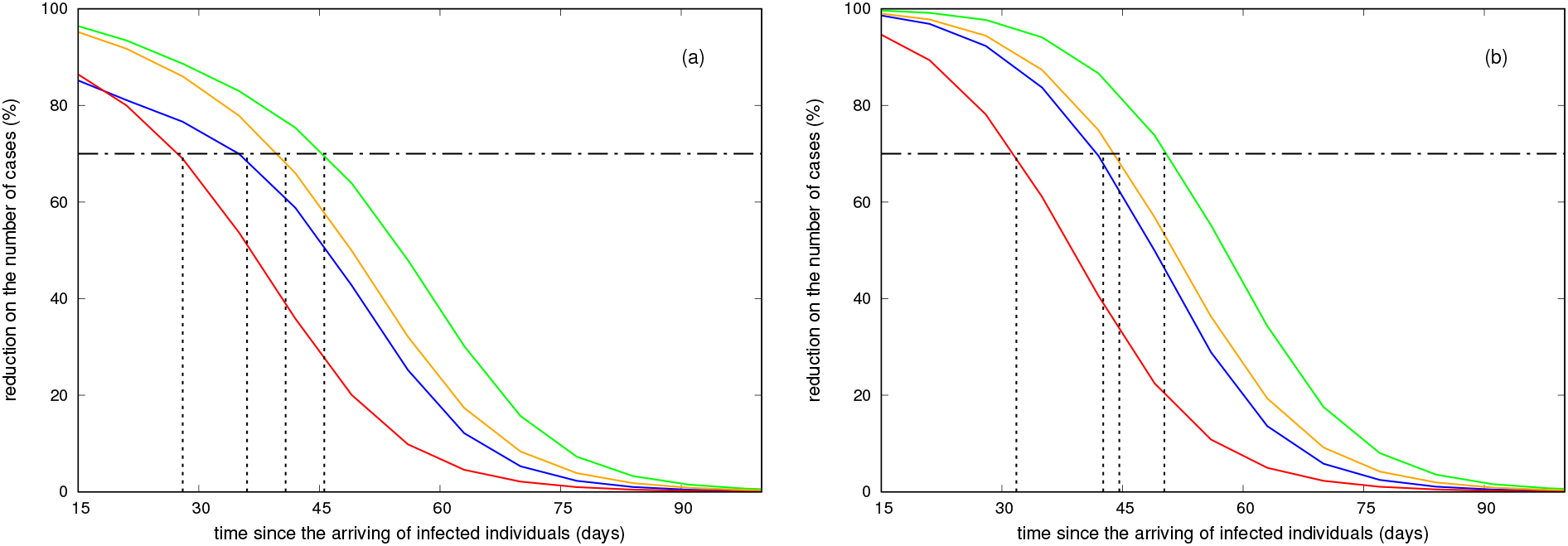
Reduction on the number of cases versus time of starting control. The parameters are the same as in Figure 4.

The partial rank correlation coefficient (PRCC) obtained from a global sensitivity analysis^15^ is shown in Figure 6. We run 3000 simulations that correspond to different input parameter sets, all of them related to control measures; the output is control efficacy.

**Figure 6.**
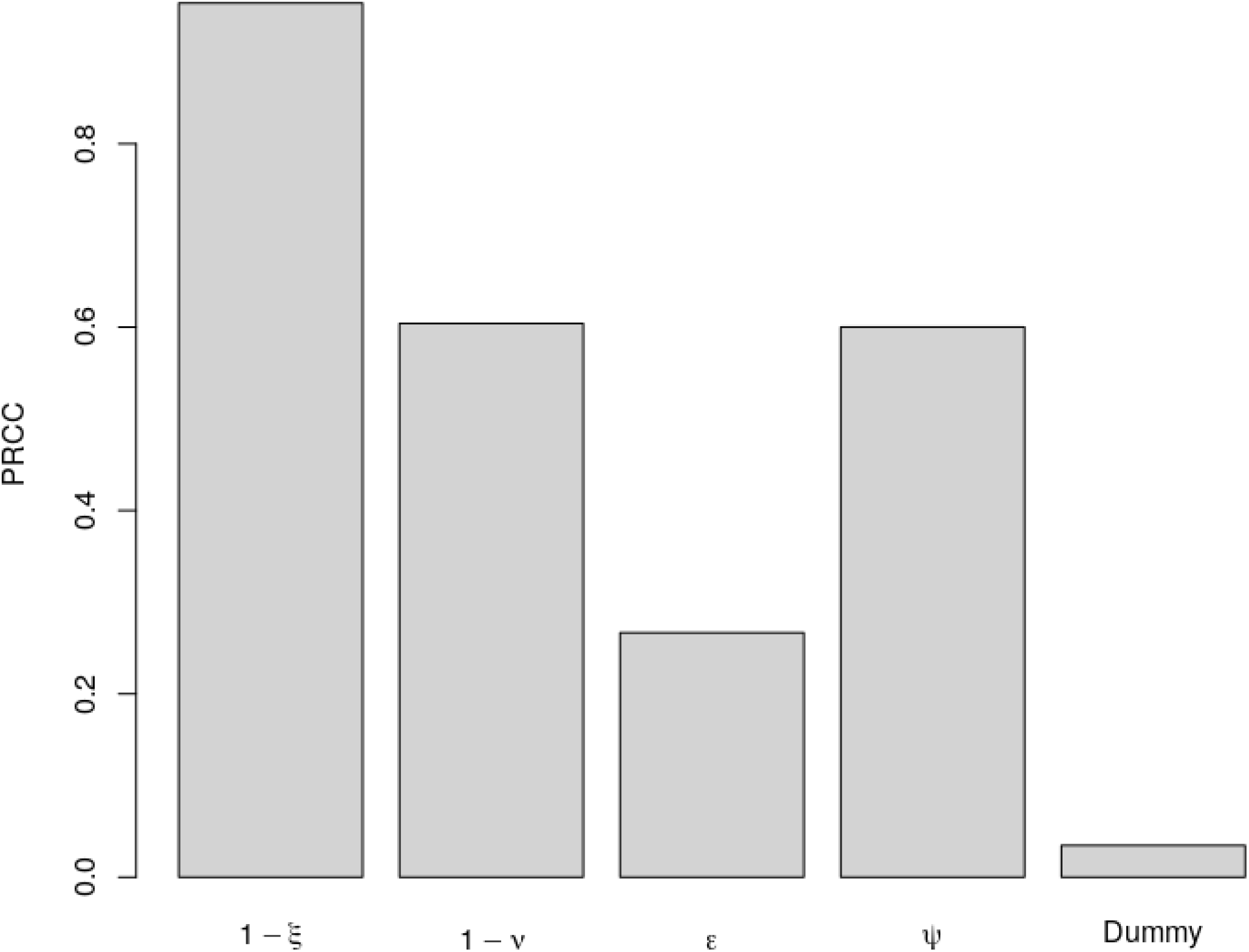
Sensitivity analysis using control efficacy as the output. A negative-control, dummy parameter, was used to assign a zero value for a sensitivity index.

Two dendrograms were obtained from clustering the municipalities and they are shown in Figure 7. The first one regarded pyramid age, temperature, and humidity. On the second one, we included population density to the list of variables. One city might belong to different clusters in each dendrogram and the differences between the dendrograms are highlighted by the gray lines connecting both.

**Figure 7.**
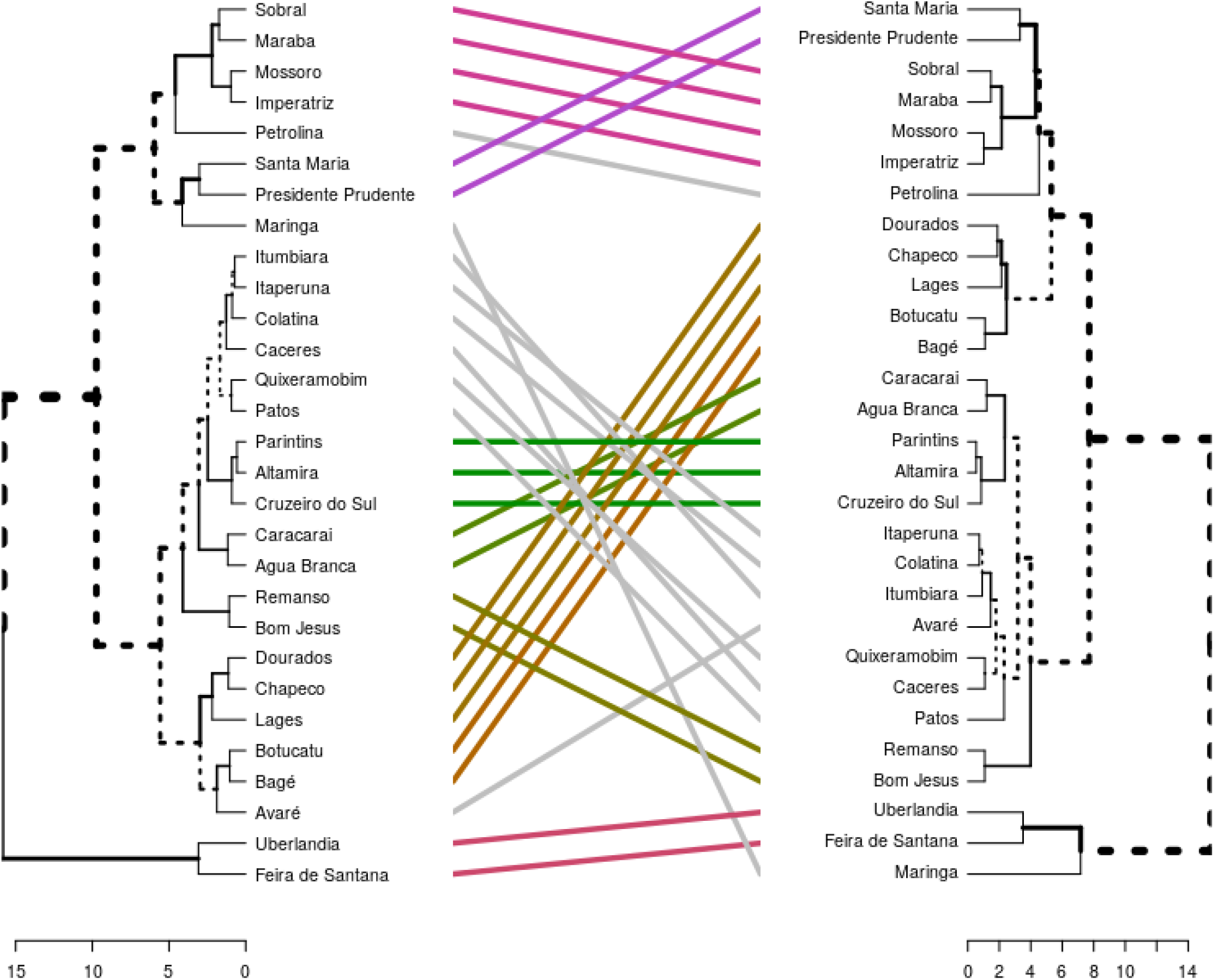
The municipalities are clustered in two ways, from left to right: (1) based on theirs age pyramid, temperature, and humidity, and (2) the same variables plus population density.

## Discussion

Ranking the cities by the number of cases (Figure 3a) we can see that, when we sum over all ages classes, the coldest weather cities (Lages, Bagé, Botucatu, Chapecó and Santa Maria) appers on the top. Botucatu and Chapecó have the same mean temperature, but as Botucatu is drier than the other, it is in a higher rank compared to Chapecó. At the other pole, the least infected cities have higher temperature and humidity: Mossoró, Caracaraí, Patos, Parintins and Marabá. The result basically reproduces the rank of *R*_0_ value of each city. The expression of *R*_0_ was obtained from studies of COVID-19 transmission on China that suggest this relationship^13^. However, as know from many other respiratory diseases like influenza, climate variables are important factors that drive the spatio-temporal behaviour of the disease transmission, but are not the only ones. Also, the contribution of each factor still be a topic of research and it looks that they do not have the same impact when considering areas of temperate or tropical weather^14^. A general consensus is that they explain about 30% of disease incidence. When studying capitals cities of states of Brazil, we see that Fortaleza-CE and Manaus-AM are some of the most affected cities by COVID-19, respectively 3482 and 2160 confirmed cases, despite being hot and wet metropolitan areas. Studies crossing health status of population with socioeconomic dimension such as education, poverty and deprivation of resources and service put the states of Amazônia and Ceará with bad index value, from -1 (the worst value of index) to 1 (the best value of the index), the municipalities that compose these two states have *−*0.77 *±* 0.12 and *−*0.63 *±*0.09, respectively^16^. Besides, the existence of an international airport in both cities, riverport and seaport, respectively, at Manaus and Fortaleza, put them in a position of right risk for the introduction and spreading of new pathogens.

When seeing the number of cumulative cases specifically on the risk group (individuals older than 50 years), we have on the top: Bagé, Santa Maria, Itaperuna, Presidente Prudente and Botucatu, and on the bottom Marabá, Cruzeiro do Sul, Caracaraí, Parintins, and Altamira. In this case, the age pyramid and the contact matrix have a great influence on ranking the cities. In particular, the ones that are on the top have approximately 25% of individuals in age classes greather than 50 years old, and the ones on the bottom about 12%. Besides, the difference on the cumulative number of cases specifically on the risk group (individuals older than 50 years) between the extreme cities of the list (Bagé and Marabá) is higher than when we compare the difference between cities considering all infected individuals cases (Lages and Mossoró).

Comparing the proportion of lethal cases (Figure 3b), we can see that when we sum over all age classes, we obtain more or less the ranking observed for the cumulative number of cases for individuals older than 50 years old, i.e., on the top Bagé, Santa Maria, Itaperuna, Presidente Prudente, and Botucatu, and on the bottom Marabá, Caracaraí, Cruzeiro do Sul, Altamira and Parintins. On the other side, summing over the classes of individuals over than 50 years old, the ranking for lethal cases are: from the bottom, Caracaraí, Marabá, Chapecó, Altamira and Dourados, and from the top, Quixeramobim, Patos, Aguá Branca, Remanso, and Bagé. For this last group, the proportion of individuals in the age class where the mortality is higher achieves almost 42%.

It is expected that any kind of control on disease transmission will affect the epidemics course, by delaying and reducing its peak. The gain on smaller numbers of infected individuals during the course of the epidemics is obtained by increasing the duration of it. Since there is no vaccine, mitigation strategies rely on social distancing, isolation of infected individuals, self-isolation when you are a suspected case, mandatory quarantine applied to all populations, and travel restrictions^17^. So, we drawn scenarios with different strategies and interventions. We can clearly see that for each city, we have an optimal control measure, depending on the target. Hypothetically, lets consider that a reduction of 60% on the number of the cases is needed to control the epidemics in all the cities represented in Figure 4. Patos would need a 50% reduction on the social contact, while Caracaraí would need almost 60%. The general characteristics of the curves are kept on both panels, but when testing more people (*ψ* = 0.3), you may achieve the same efficacy while decreasing social distancing.

The effect of delaying the start of controls were modelled as well. The result shows, again, a specific behavior for each municipality, but they all have in common one fact: the earlier cities start control measures, the greater is the reduction on the number of the cases. Testing more people in the first 30 days is surely the best choice and testing more people may also allow delaying social distance; for 70% of efficacy we obtained in mean 5 days.

The sensitivity analysis order the importance of parameters on control efficacy which is (decreasing order) *ξ, {ν, ψ}* and *ε*, highlighting the importance of mandatory quarantine and testing individuals for COVID-19. Combining isolation of detected COVID-19 positive cases with social distancing can provide an efficient way of halting or diminishing disease incidence on population, but the control effort will depend on characteristic of each municipality.

We sustain the hypothesis that each city must be individually studied, but when we compare them, we can cluster and show patterns. This emphasize the importance of studying the impact of climatic, demographic, cultural and social variables on disease dynamics. Maringá, Lages, Caracaraí, and Patos belong to different groups that differ by the age structure, temperature, humidity, and population density. Maringá is a big city, with moderate temperature and humidity, and a considerable amount of elderlies. Lages and Patos are cities with cold weather and hot weather, respectively, and with median size and pyramid age homogenous distributed. Lastly, Caracaraí is a small city, with soft weather and a younger population. We consider that they are very representative of what we can find in an inner Brazil.

In huge and heterogeneous countries like Brazil, we expect that many factors, such as population density, temperature and mobility will modulate disease transmission. Quantifying and identifying such contributions can help governments to take decisions about mitigation strategies. The knowledge about other respiratory infection disease that assault the population in different parts of Brazil such as influenza can provide a pool of important information that may be useful to forecasting COVID-19 in many municipalities.

This is the first paper describing mathematical models for small and medium cities of inner Brazil. We show here that different control measures should be taken for different cities and, most important, each city may have an optimal combination of general social distance with testing and isolating positive cases that control the epidemics curve and permit the health systems to be prepared for the peak of the number of cases. By comparing cities and models, we suggest patterns of the evolution of number of cases and control strategies. As testing is a major issue for great part of nations at this moment of the pandemic, social distance in different degrees should be established.

## Methods

### Municipalities

We aimed a study capable of representing most of the small and medium cities of Brazil. Therefore, we decided to choose municipalities from different states and regions, with varied population density, temperature, humidity, as well as age structured. From North we have: Altamira-PA, Marabá-PA, Cruzeiro do Sul-AC, Parintins-AM, Caracaraí-RR; from Northeast: Agua Branca-AL, Sobral-CE, Quixeramobim-CE, Bom Jesus-PI, Imperatriz-MA, Mossoró-RN, Patos-PB, Petrolina-PE, Feira de Santana-BA, Remanso-BA; from Central-West: Dourados-MT, Caceres-MT, Itumbiara-GO; from South: Santa Maria-RS, Bagé-RS, Lages-SC, Chapecó-SC, Maringá-PR; and from Southest: Uberlândia-MG, Avaré-SP, Botucatu-SP, Colatina-ES, Itaperuan-RJ, Presidente Prudente-SP. Figure 1 shows the geographic localization of each one, into Brazil map. Table 1 summarizes information about each city that can modulate Covid-19 transmission. In particular, temperature and humidity corresponds to the mean values in April month^18^.

After listing the cities, we clustered them. We first grouped cities by age pyramid, temperature and humidity, which, in association with total populational size, may be the most important characteristics that modulate the epidemic curve. Afterwards, we added population density and clustered all cities again. We used a hierarchical agglomerative clustering method, combining cluster thrown the complete linkage criterion and Euclidean distance as a metric to measure dissimilarity between the observation sets.

By taking each city as a model, studying the main characteristics, and crossing into a cluster study, we believe its possible to extrapolate the results of this study to other cities that won’t be plotted here. Similar socioeconomics and climate characteristics should give analogous results.

### Mathematical model

The proposed model is an age structured one that divides the human population into fifteen age groups: 0 to 4 years, 5 years interval from 5 to 70 years, and greather that 70 years^19^. The variables of the model are *t, S*_*i*_ := *S*_*i*_(*t*), *E*_*i*_ := *E*_*i*_(*t*), *I*_*i*_ := *I*_*i*_(*t*), *Q*_*i*_ := *Q*_*i*_(*t*), *R*_*i*_ := *R*_*i*_(*t*); respectively, time, susceptibles, exposed, infected, detected and isolated infected individuals, and recovered one. The index *i* is the age class. The natural mortality rate *µ* appers in all age classes, and from 1 to 15, the parameter *α*_*i*_ takes into account the transition among them. Individuals are born susceptible, and they become exposed, when contacting infected or isolated individuals at rate *β*_1_ and *β*_2_ = *νβ*_1_ (*ν∈* [0, 1]), respectively. The parameter *c*_*i,j*_ represents the fraction of daily contacts that age group *i* has with age group *j*. Target control can be done by varing *ξ*_*i*_ *∈* [0, 1], being *ξ*_*i*_ = 0 complete protection of class *i* and *ξ*_*i*_ = 1 no protection of class *i* against the infection. After a period of time *η*^*−*1^ exposed individuals becomes infectious. At rate *ε*, a fraction *ψ ∈* [0, 1] of infected individuals are identified and isolated. Additional mortality related to the disease is considered in the compartiments of infected and isolated individuals, *σ*_*i*_. Finally, these individuals become recovered at rates *γ* and *τ*. The ODE model is given by

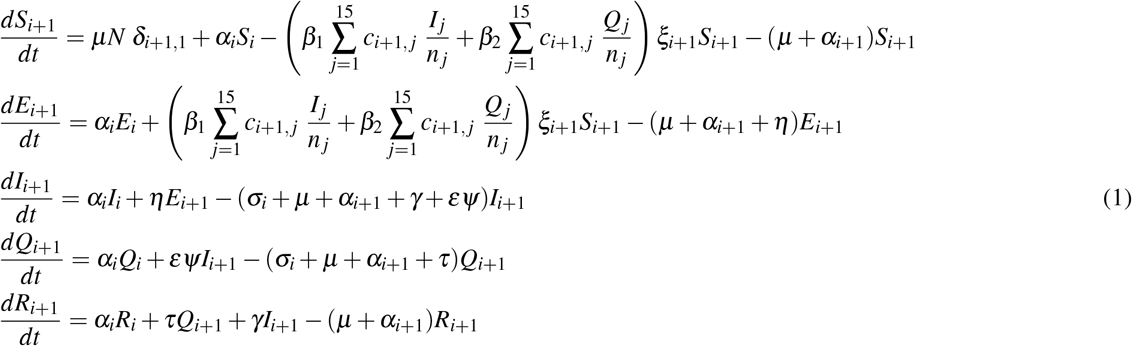

with *i* = 0, …, 14, *α*_0_ = *α*_15_ = 0, *α*_1_ = … = *α*_14_ = *α, δ*_1,1_ = 1, and *δ* _*j*,1_ = 0 with *j* ≠ 1. Besides, *n* _*j*_ = *S* _*j*_ + *E*_*j*_ + *I*_*j*_ + *Q*_*j*_ + *R*_*j*_, and 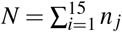 at *t* = 0. Table 2 summarizes model parameters, their description, range of values and units. Figure 2 shows the diagram of the compartmental model.

Defining 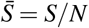 we can rewrite (1) as

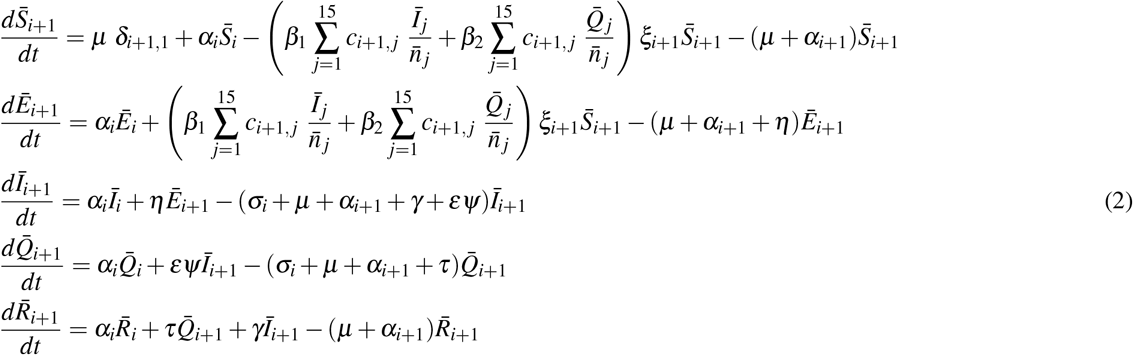

with 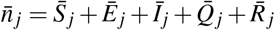 The disease free equilibrium is given by

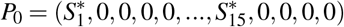

Where

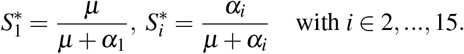

In order to obtain the next generation matrix^20^,21, we used the reduced system given, in its vectorial form, by

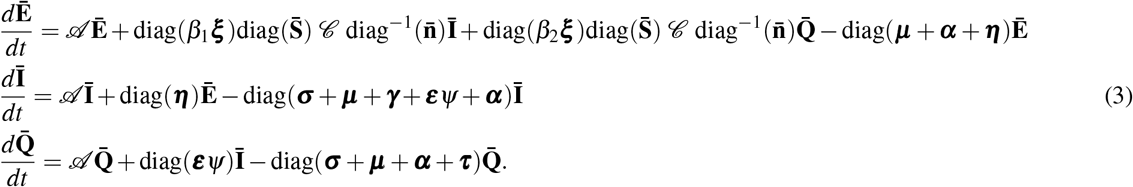

Bold symbols represent vectors as ***x*** = [*x*_1_, …, *x*_15_]^*T*^ and diag(***x***) represent diagonal matrices, *M* = [*m*_*i j*_], in which *m*_*ii*_ = *x*_*i*_. 𝒷 is the contact-distribution matrix among the age groups^22^, and

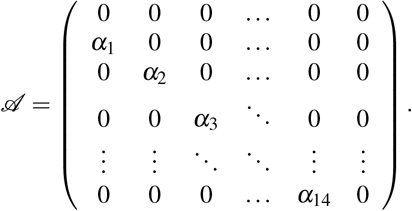

The matrix of infection terms, ℱ, and the matrix of transition terms, 𝒱, are given, respectively, by

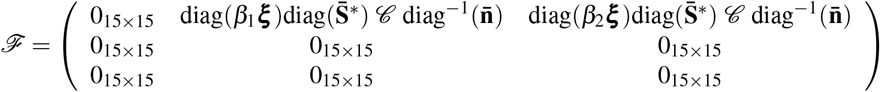

and

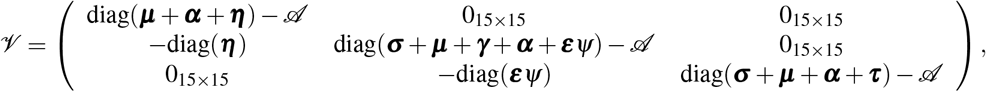

in which 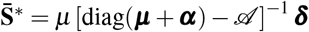 with ***δ*** = [1, 0, …, 0]^*T*^, is the disease-free equilibrium of (2). The basic reproductive number denoted by *R*_0_ is given by the spectral radius of the next generator operator matrix given by ℱ𝒱 ^*−*1^ (i.e. its dominant eigenvalue). The disease-free equilibrium 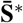 is locally asymptotically stable if *R*_0_ *<* 1, and unstable if *R*_0_ *>* 1. *R*_0_ is the mean number of secondary cases that a primary case generates in a whole susceptible population, which implies before control measures. A simple and direct way to calculate the effort to be done to control an epidemic is given by *P*_*c*_ = 1 *−* 1*/R*_0_, where *P*_*c*_ is the fraction of population that likely to be infected without mitigation. This represents the worst scenario since the deterministic approach has several assumptions like large population, well-mixed individuals, and no spatial structure.

### Simulations

In all simulations, the parameter *β*_1_ was calibrated using the parameters given in Table 2 (or specified in the text) and the next generation matrix, under no control measure. The addition mortality rates (days^*−*1^) are calculated through the expression

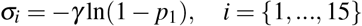

where *p*_*i*_ is the probability that an individual at age group *i* dies during his infectious period. We have *p*_1_ = *p*_2_ = 0, *p*_3_ = … = *p*_8_ = 0.002, *p*_9_ = *p*_10_ = 0.004, *p*_11_ = *p*_12_ = 0.013, *p*_13_ = *p*_14_ = 0.036, and *p*_15_ is weighted by the number of individuals in age group with more than 80 years old (*x*_1_) and in the age group between 75 to 79 years old (*x*_2_)^10^,23 giving

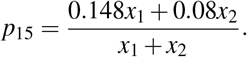

The simulations start with ten infected individuals (in the age class of 25 to 50 years) introduced in a whole susceptible population. Control started later, after one month since the introduction of infected individuals.

Control was explored by reducing contact rate among age classes (*ξ*), decreasing the time of detection of infected individuals (*ε*^*−*1^), increasing the fraction of individuals that are detected and isolated (*ψ*), and decreasing the contribution of detected and isolated individuals to the disease transmission (*ν*). A connectivity-based clustering was used to group municipalities by their similarities.

Two different scenarious were analysed. The first one deals with a situation where the detection and isolation of infected individuals occurs in a short period of time. Therefore, we set up *ε*^*−*1^ to 1 and 2 days and *τ*^*−*1^ = 6 (*≈γ*^*−*1^) days. The second one suppose that detection tooks long time, then *ε*^*−*1^ (*≈ γ*^*−*1^) was set up to 5 and 6 days and *τ*^*−*1^ = 2 days. The other parameters are *β*_2_ = 0.55*β*_1_ days^*−*1^, *η*^*−*1^ = 3 days, *γ*^*−*1^ = 6.4 days, and *µ* = 3.65 *×* 10^*−*5^ days. In general, the Figures were done with the set of parameters that represent the late detection.

Since the time of starting control impacts the evolution of disease transmission, the efficacy of control was measured varying this parameter in the simulation. For this, we measure the reduction (in percentage) on the number of infected individuals with and without control. Target and no target control over higher age classes was explored by ranking and comparing the municipalities by the acumulative number of infected individuals, and by the proportion of lethal cases. Finally, a sensitivity analysis based on partial rank correlation coefficient (PRCC) was done to discuss the contribution of each model control parameter to the control efficacy, measured as the percentage of infected cases that are avoided. The input parameters were *ε, ξ, ν*, and *ψ*. The first one took from a uniform distribution from 0.166 to 0.2 (late detection) and from 0.5 to 1 (early detection), and the others one from an uniform distribution in the range of 0 to 1.

## Data Availability

All data used are avaible on line, on scientific database

## Acknowledgement

T.N.V. and C.P.F. thanks support from São Paulo Research Foundation (FAPESP) grant 18/24811-1 and 18/24058-1.

## Author contributions statement

C.M.C.B.F. and G.B.de A. conceived the problem, T.N.V and C.P.F conduct the simulations. All authors analysed the results and reviewed the manuscript.

## Additional information

### Competing interests

We declare no competing interests.

## Notes

### Competing Interest Statement

The authors have declared no competing interest.

### Funding Statement

NO external funding was received for this work

